# A Retrospective Study to Determine the Impact of Psychedelic Therapy for Dimensional Measures of Wellness: A Quantitative Analysis

**DOI:** 10.1101/2023.05.10.23289787

**Authors:** Victoria Di Virgilio, Amir Minerbi, Jagpaul Kaur Deol, Salena Aggerwal, Toufik Safi, Gaurav Gupta

## Abstract

**Background:** The World Health Organization (WHO) defines wellness as the optimal state of health of individuals and groups. No study to date has identified the impact of psychedelic medicines on optimizing wellness using a dimensional approach. Using this approach, treatment effects can be measured more broadly using a composite score of participants’ global perceptions of change for pain, function, and mood scores. Given the precedence in previous work for retrospective studies of participants’ self-medicating with these substances, the nature of this study design allows for a safe way to develop further evidence in this area of care, with wellness as the broad indication.

**Methods:** 65 civilian or military veterans above the age of 18, self-identifying as having used psychedelic medicines for non-recreational purposes in the last 3 years were recruited. Participants completed the following standardized questionnaires: Patient Global Impression of Change (PGIC) scale, Pain, Enjoyment of Life and General Activity (PEG) scale, Anxiety and Depression scale (ADS), and Disability Index (DI) scale. The analysis focused on reported PGIC outcomes and correlations between subscales. Given the nature of the study, a comparison to the baseline could not be made.

**Results:** On average, participants reported improvement in all domains (pain, mental health, function, and overall quality of life), regardless of the medicine. Perceived improvement was highest in mental health and overall quality of life, and lowest in pain. Kendall correlation showed a highly significant association between the perceived changes in all domains. Correlation coefficients were highest between the perceived change in function, quality of life, and mental health.

**Discussion:** The use of various psychedelic medicines may be associated with a broad range of changes that could help clarify the mechanism of how they impact wellness in the future. Pain, mental health, function, and overall quality of life accordingly improved after the use of these medicines. Minor differences between the drugs were not found as significant, indicating that the perceived benefits seemed to be specific to the psychedelic class. Numerous limitations exist to this type of study which was relatively small in size, retrospective and anonymous in nature.

**Conclusion:** The wellness of individuals or groups is not simply an absence of disease, symptoms, or impairments. Instead, it is an outcome that is shaped by a myriad of personal characteristics, psychophysiology, and choices, expressed throughout one’s lifespan, unfolding in dynamic interaction with a complicated sociocultural and physical environment.

## Background

The World Health Organization (WHO) constitution states: “*Health is a state of complete physical, mental, and social well-being and not merely the absence of disease or infirmity*.” This definition has a significant implication that health is not merely the absence of diseases or physical and mental health conditions, but rather is a comprehensive state of well-being. It encompasses an individual’s awareness of their own capabilities, their ability to handle everyday challenges, ability to function effectively, and ability to give back to their community (*Health and Wellbeing*, n.d.).

Moreover, research regarding the Diagnostic and Statistical Manuals (DSM) guidelines, shows that diagnostic criteria are not extremely specific, inter-rater reliability is low, and treatment outcomes are not always correlated with clinical indicators (Bremmer et al., 2008; Hahn et al., 2010; Lacasse & Leo, 2005; Lieblich et al., 2015; Liu et al., 2015; Maas et al., 2009; Osimo et al., 2020). In addition, mental health treatments are being employed for clusters of mental health symptoms, not necessarily disorders. For example, numerous more individuals have been prescribed antidepressant medications than those who received a diagnosis of depression (Moncrieff, 2018; Whitaker, 2005). As a result, many authors advocate for a dimensional approach that considers symptomatology, severity, life impact, and treatment response rather than diagnostic categories (Brown & Barlow, 2005; Coghill & Sonuga-Barke, 2012; Widiger & Samuel, 2005).

Therefore combining a dimensional approach with emerging therapies like psychedelic medicines could be an effective validation method. Individuals with varying degrees of symptomatology could then be compared regardless of their underlying clinical diagnoses (Di Virgilio et al., 2023). This would build on recent studies about psychedelics for different mental health indications supporting benefits for various conditions, such as addiction, mood, end-of-life care, and pain and anxiety disorders (Bornemann et al., 2021; Gasser et al., 2015; Malone et al., 2018; Nielson et al., 2018; Watts et al., 2017). Prior studies have suggested that these treatments could potentially yield clinical benefits, such as reduced severity of anxiety/mood/pain symptoms, long-term clinical effects without maintenance therapy, less reliance on cannabis and possible reduced rates of substance abuse and relapse (Elsey, 2017; Herzog et al., 2018; Nielson et al., 2018; Schlag et al., 2021; Tupper et al., 2015). Additional benefits could include bolstered prosocial behavior, reduced public healthcare utilization and improved reintegration from personal, community, occupational standpoints (Elsey, 2017; Herzog et al., 2018; Nielson et al., 2018; Schlag et al., 2021; Tupper et al., 2015).

However, various limitations to these studies exist (Elsey, 2017; Herzog et al., 2018; Pratt et al., 2019; Tupper et al., 2015). This includes the selection of participants using categorical/diagnostic conditions since this limits the ability to extrapolate outcomes to real-world scenarios because patients are often affected by more than one health problem or do not meet the strict criteria for any one disease. Furthermore, as all psychedelic medicines are used in similar ways, across indications and appear to be safe, it would be useful to study them comparatively using a dimensional approach for overall well-being. Using this approach employed in our study, we believe could lead to a richer understanding of the degree and scope of benefit that these medicines offer (Bornemann et al., 2021; Castellanos et al., 2020; Di Virgilio et al., 2023; Tupper et al., 2015).

We therefore designed this study to measure the treatment effects more broadly using a composite score of participant global perception of change, along with pain, function, and mood scores, which has not been done to date (Di Virgilio et al., 2023). The other part of this study involved the qualitative analysis of the comments left by the participants (Di Virgilio et al., 2023) using this approach. These comments were categorized into various themes and described improvements and experiences for this class of medicines. The quantitative outcomes of this dataset will be reported here and may further contextualize the effect of these medications on improving wellness. (Di Virgilio et al., 2023).

## Methods

Given the previous precedence for retrospective study of participants self-medicating with psychedelics, this study design allows for a safe way to develop further evidence in this area of care (Bornemann et al., 2021; Schindler et al., 2015). The clinical trial registration number is NCT05469243.

65 participants self-identifying as having used psychedelic medicine for non-recreational purposes were recruited for this study. To be considered eligible for admission to the study, they were civilian or military veterans between the ages of 18-99 with a self-reported past of psychedelic medicine used for therapeutic purposes in the last 3 years. Self-reported past psychedelic medicine use for recreational purposes and by active military members were exclusions for participation.

Participants completed the following standardized psychometric measures: Patient Global Impression of Change (PGIC) scale measured for overall quality of life, anxiety, mood, pain and disability subscales, the Pain, Enjoyment of Life and General Activity (PEG) scale, the Anxiety and Depression scale (ADS), and the Disability Index (DI). Participants also provided demographic information, and information on their medical history, nature/indications for use (i.e. depression, PTSD, chronic pain, other), and adverse events. The scores are expressed based on how they felt when reporting concerning the numerous dimensions of wellness. No change in scores was measured, but those reaching a score of 5 on the PGIC scale were considered clinically improved.

### Statistical analysis

Dichotomous variables were compared using the Chi-Square test. Quantitative variables were evaluated for normality of distribution using the Wilk-Shapiro test. Normally distributed variables were compared using ANOVA while non-normally distributed variables were compared using the non-parametric Kruskal-Wallis test. Adjustment for multiple comparisons was performed using Benjamini-Hochberg correction. Correlation analysis was done using the non-parametric Kendall Tau test. Analyses were done on IBM SPSS version 28.

## Results

Sixty-five participants agreed to participate in the study and filled in the study questionnaires. Participants were predominantly women (58%), and the mean age was 46±13.5 years. Participants were grouped based on the psychedelic drug they reported using most recently into four groups: psilocybin (24 participants), ayahuasca (8 participants), ketamine (5 participants), amphetamines including MDMA (4 participants), and others which includes ibogaine (2), LSD (2), cannabis (4), other medicines (4), and not specified (11). Participants in the five groups were comparable in their demographic measures, self-reported mental health history, and their scores in pain, disability, anxiety, and depression questionnaires (Table 1).

**Table 1:**
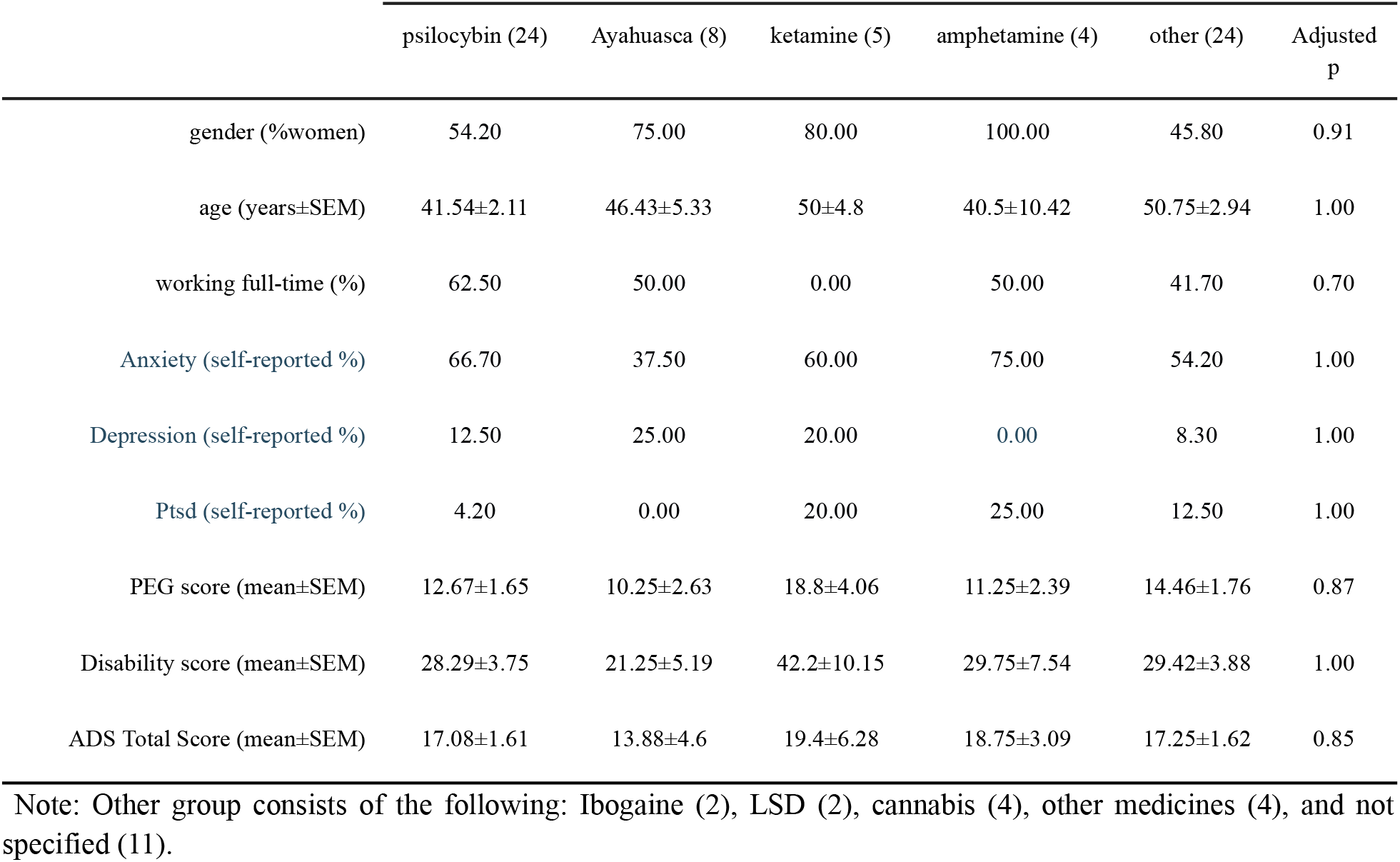
Demographic properties, and questionnaire scores of study participants.

Participants reported on the perceived effect of taking psychedelic drugs in several domains: pain, mental health, function, and overall quality of life. On average, participants reported improvement in all domains, regardless of the drug they were taking. Perceived improvement was highest in mental health and overall quality of life and lowest in pain (Figure 1). When comparing the differences in the perceived change between patients who took different drugs, small differences were observed: participants taking ketamine reported smaller improvement in function and mental health compared to participants taking other psychedelics. These differences were marginally significant; however when corrected for multiple comparisons, no significant changes were observed (Table 2).

**Table 2:**
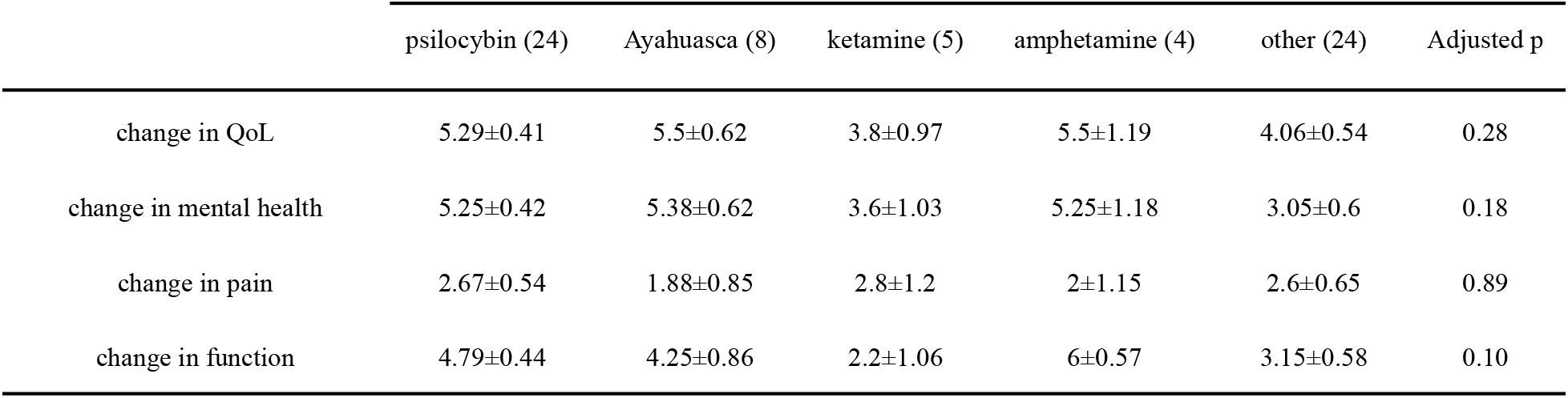
Perceived global impression of change in four domains. (QoL – quality of life)

**Figure 1.**
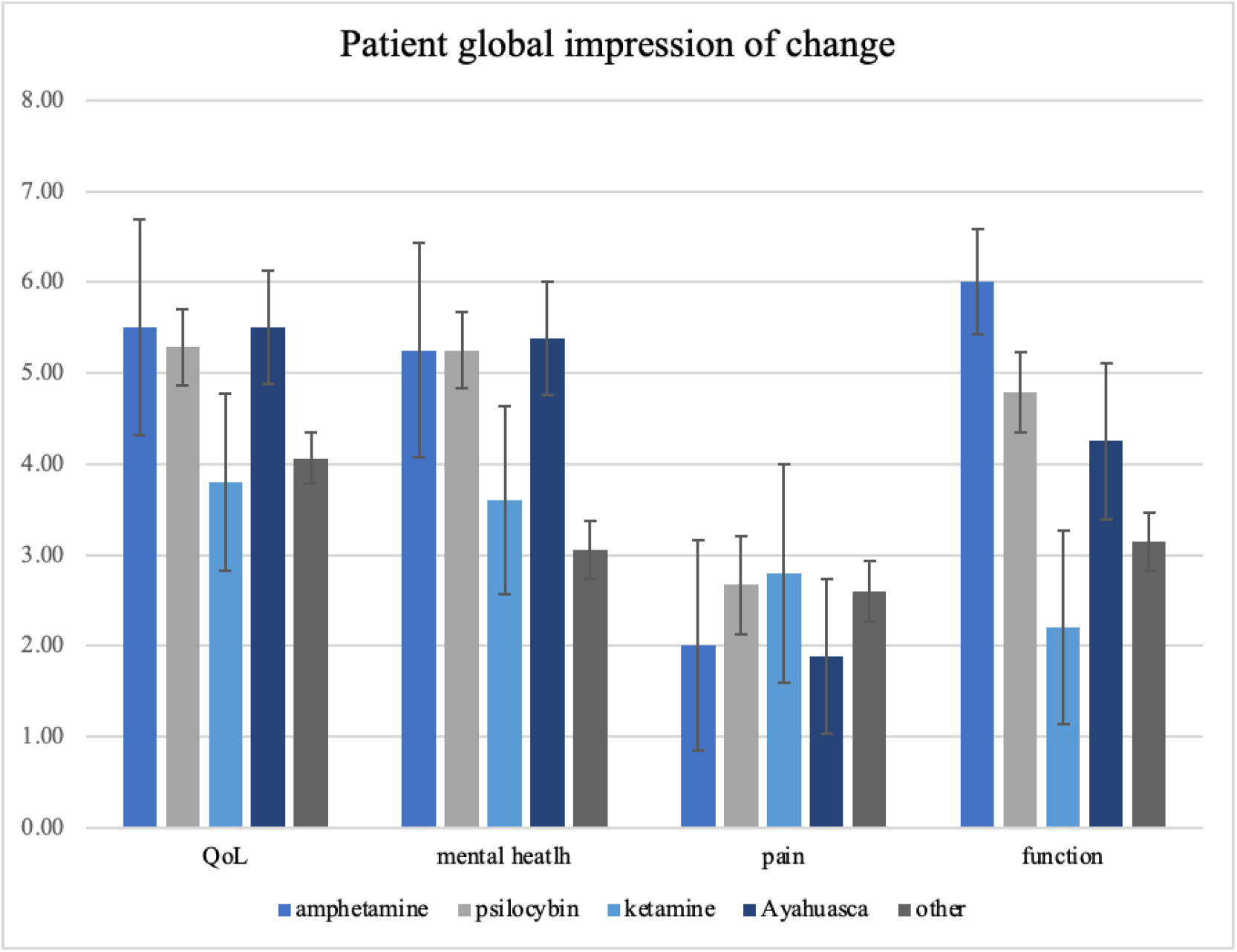
: Participant-reported perceived global impression of change following psychedelic consumption in four symptomatic domains. (Error bars indicate the standard error of the mean; QoL – quality of life).

Next, we investigated the association between the perceived change in the various domains. Kendall correlation showed a highly significant association between the perceived changes in all domains. Correlation coefficients were highest between the perceived change in function, quality of life, and mental health (Table 3).

**Table 3:** Kendall correlation analysis of perceived global impression of change questionnaire scores. (*p<0.05; **p>0.01).

|  | change in QoL | change in mental health | change in pain | change in function |
| --- | --- | --- | --- | --- |
| change in QoL | 1.00 | 0.79** | 0.31** | 0.72** |
| change in mental health | 0.79** | 1.00 | 0.29** | 0.73** |
| change in pain | 0.31** | 0.29** | 1.00 | 0.35** |
| change in function | 0.72** | 0.73** | 0.35** | 1.00 |

Also analyzed was where people fell on average for the following patient-reported outcomes: the Pain, Enjoyment of Life, and General Activity (PEG) scale, the Anxiety and Depression scale (ADS), and the Disability Index (DI) (Table 1). These measures quantify the participants’ pain, mental health, and function and describe, based on how the participants felt at the time of reporting (i.e not change scores). Depending on the medication, participants fell into the moderate range for the PEG scale (30-70%), into the severe range for the total ADS scale (11-21), and the mild to moderate range for the DI (20-50%).

## Discussion

The various dimensions underlying health and wellness are embedded across diagnostic criteria employed by classic categorical systems and thus are frequently overlooked in medicine, public policy, and industry. Treatment outcomes using this approach can be complicated to differentiate due to significant comorbidities between various mental and physical conditions, as participants afflicted with comorbid conditions are often not included in studies (Cucinello-Ragland & Edwards, 2021; Kirsh, 2010; Kleykamp et al., 2021; Kohrt et al., 2018; McParland et al., 2021; Myhr & Augestad, 2013; Weeks et al., 2016). To further explain the underpinning of these challenges some authors highlight overlapping symptomatology, treatments, and functional consequences, while other authors argue causative relationships where these conditions influence each other (Aronoff, 2016; Di Virgilio et al., 2023; Fornasari, 2012; Kynast et al., 2013; Rieder et al., 2017).

However, there is a modern movement towards precision-based healthcare centered on the individual pursuit of “wellness”, regardless if that individual meets specific diagnostic criteria (Dawson, 1989; Kraft & Goodell, 1993). The long-term goal of this type of research would be to determine what factors about the participant selection and treatment can conceivably be changed to facilitate outcomes (Belser et al., 2017). This is particularly important when examining the outcomes of emerging fields like the clinical applications of psychedelic medicines. Although there is some early optimism regarding treatment with psychedelic medicines, there are still many details to consider such as indications, contraindications, efficacy compared to similar treatments, and cost benefits.

Until now, there has not been a study to date that has looked at the treatment of wellness with psychedelics using measures that will yield generalizable outcomes. The data from this study showed that pain, mental health, function, and overall quality of life improved after using these medicines. Small differences between the drugs were not found as significant, indicating that the perceived benefits seemed to be specific to the psychedelic class. The use of various psychedelic medicines appears to be associated with a broad range of changes that could help clarify the mechanism of how they impact wellness in the future.

As noted, perceived benefits appear to be specific to the psychedelic class, which was initially noted in the qualitative analysis of this study (Di Virgilio et al., 2023). Small differences in categorical improvement were found between the medicines, but after adjustments the differences between the drugs were not found as significant, indicating that the benefits are class specific. Understanding the improvement in wellness from psychedelic medicines might be useful from a clinical point of view and improve the limited access for medical and mental health professionals.

The Pain, Enjoyment of Life, and General Activity (PEG) scale, the Anxiety and Depression scale (ADS), and the Disability Index (DI) measures quantified how the participants felt at the time of reporting in terms of pain, mental health, and function. Based on this data, it is estimated that before participants’ psychedelic medicine use, these measures were on average higher, since the impression of change scores were improved, however, no change in scores was collected. In future collecting baseline and follow up scores and correlating with wellness scores and subscales will be imperative (Baernholdt et al., 2012). Nonetheless, the scores themselves are still elevated suggesting most patients do not improve fully, and could help explain the full time work rates in this middle aged cohort.

Generally, the research for psychedelic medicines is promising for treating various mental health disorders and promoting wellness in the general population (Rootman et al., 2021). A previous review study suggested that psilocybin-assisted psychotherapy may provide a therapeutic advantage over other treatments such as transcranial magnetic stimulation, electroconvulsive therapy, or ketamine infusion therapy for various mental health disorders (Reiff et al., 2020). Additionally, research involving psilocybin-assisted psychotherapy for the treatment of major depressive disorder showed symptomatology decreases in comparison to baseline scores for a year following treatment (Gukasyan et al., 2022). Finally, psychedelic medicine micro-dosing in an international sample of participants with and without diagnosed mental health conditions displays how these medicines may be useful for the promotion of wellness (Rootman et al., 2021). Therefore our results, in general, are consistent with previous studies.

In terms of benefits these medicines may be associated with broad clinical application and durability, although more research is required. Other advantages compared to “standard” treatments include fewer treatment visits, easier accessibility where regulations permit, and long-term cost (Mastikhina et al., 2020; Reiff et al., 2020). For these reasons, further studies to analyze the mechanisms of psychedelic medicines that lead to quantitative and qualitative improvements in wellness are required. Furthermore, investigating the relationship between the neurobiology of the medicine and the patient will improve the precision of wellness applications (Di Virgilio et al., 2023; Nielson et al., 2018).

Limitations of this study include the retrospective nature and sample size. The results were based on self-report, with the multiple measurements to indicate changes in patient-reported outcomes. The outcome measures will also require further standardization. Perceived change over time was not analyzed and there was no inclusion of a control group. Finally, understanding the neurophysiological effect will be key in treatment planning and personalization of care. Future studies should work toward a deeper understanding of these effects, and the potential for psychedelic medicines to support and maintain wellness throughout the lifespan of a patient.

## Conclusion

Pain, mental health, function, and overall quality of life consequently improved after the use of these medicines. Small differences between the drugs were not found as significant, indicating that the perceived benefits seemed to be specific to the psychedelic class. The perceived change in these dimensions were statistically significant, indicating that the standard measures to evaluate pain, mental health, and functioning scores have improved over time. However, it would be beneficial to verify these findings in future studies. The use of various psychedelic medicines appears to be associated with a broad range of changes that could help clarify the mechanism of how they impact wellness in the future.

## Data Availability

All data produced in the present work are contained in the manuscript

